# Novel document-level measures and their performance for active learning uncertainty sampling—a use case for automatic CDSS ontology curation

**DOI:** 10.1101/2025.04.15.25325868

**Authors:** Shailesh Alluri, Keerthana Komatineni, Rohan Goli, Richard D. Boyce, Nina Hubig, Hua Min, Yang Gong, Dean F. Sittig, David Robinson, Paul G. Biondich, Adam Wright, Christian G. Nøhr, Timothy D. Law, Arild Faxvaag, Ronald W. Gimbel, Lior Rennert, Xia Jing

## Abstract

Human-in-the-loop active learning (AL) can identify key phrases to facilitate biomedical ontology development, maintenance, and other curation tasks. However, determining which documents to annotate by humans is not straightforward. We explored new strategies to make the document selection process transparent, reproducible, and effective. Our AL pipeline modified a BiLSTM-CRF model using PubMed abstracts. We tested four novel document-level uncertainty aggregation strategies—**KPSum, KPAvg, DocSum**, and **DocAvg**—that operate over standard token-level uncertainty scores: Minimum Token Probability (MTP), Token Entropy (TE), and Margin. All strategies show significant improvement in early active learning cycles (θ_0_ to θ2) for recall and F1. The systematic evaluations show that **KPSum** (actual order) shows consistent improvement in both recall and F1. The weighted F1 (β = 5, 10) provided complementary results to raw recall and F1 (β = 1). Our work advances uncertainty sampling by introducing document-level uncertainty aggregation and shows promise in automating ontology curation.

## Introduction

Clinical Decision Support Systems (CDSS) have been a cornerstone in modern healthcare, aiding medical practitioners by offering data-driven recommendations to improve patient care^1^. Ontology is an enabling technology of the Semantic Web technologies and plays a critical role in achieving information interoperability^2^. A CDSS ontology defines the CDSS entities and their relationships. It can be crucial for standardizing clinical vocabulary and ensuring information interoperability across different healthcare systems^3^. However, building and maintaining ontologies is labor-intensive, time-consuming, and requires domain expertise. Moreover, ontologies require continuous updates to remain relevant over time. As medical knowledge rapidly evolves, manually maintaining ontologies becomes increasingly challenging. Although description logics^4^ can address some of these challenges, automating certain steps can significantly ease the overall burden of ontology development and maintenance. This paper proposes an active learning-based pipeline to automate identifying key phrases in CDSS publications. Key phrases may be single words, nouns, verbs, or phrases, and experts can evaluate them for inclusion in the CDSS ontology. Our approach leverages deep learning models and is refined by incorporating iterative human domain expert (HDE) feedback. This reduces reliance on manual selection alone while ensuring that the ontology remains accurate and current. The approach is particularly important in long-term ontology maintenance. Our pipeline prioritizes instances where the model is uncertain or where consensus is mostly needed.

## Relevant literature

Identifying key phrases in texts is a typical task in natural language processing (NLP), a critical foundation for downstream applications, e.g., ontology curation or text summarization. Traditional methods, including rule-based and statistical approaches like Term Frequency-Inverse Document Frequency and Rapid Automatic Keyword Extraction^6^, have been widely adopted for key phrase extraction. However, these methods often fail to generalize across domains, prompting the adoption of machine learning-based models, such as conditional random fields (CRFs)^7^ and sequence labeling models, for named entity recognition (NER) and key phrase identification. Models such as Bidirectional Long Short-Term Memory Network-CRF (BiLSTM-CRF)^8^ and transformer-based architectures like Bidirectional Encoder Representations from Transformers (BERT)^9^ have set new benchmarks in NER tasks. However, BiLSTM is a lightweight and interpretable model compared to BERT. For example, Alzaidy et al. utilized^10^ a BiLSTM-CRF model for key phrase identification, demonstrating the effects of hierarchical context and transfer learning for domain-specific applications. Active learning (AL) is a subfield of machine learning that aims to achieve high model performance with minimal labeled data by strategically selecting the most informative samples for annotation^11^. Settles^12^ reviewed AL strategies, uncertainty sampling, query-by-committee, and expected error reduction as key techniques. Shen, et al.^13^ employed uncertainty sampling with deep learning models, effectively reducing the labeling effort. Similarly, Siddhant and Lipton^14^ explored AL in low-resource NER tasks, demonstrating the benefits of selectively annotating high-value samples. Caufield et al. explored using large language models (GPT-3.5 and GPT-4) with structured prompts to populate ontologies to achieve similar goals^15^. Radmard, et al.^16^ utilized AL to iteratively refine NER models, focusing on identifying ambiguous or novel terms in unstructured text. Margatina, et al.^17^ extended AL techniques to transformer models, achieving significant performance gains with reduced annotation costs.

Our work differs from the above-mentioned work in two ways. First, prior studies rely solely on a small, manually labeled seed set; our approach begins with a synthetically labeled training set, enabling the model to adapt progressively to real-world patterns, as described in a prior paper^10^. Second, we introduce novel ***document-level uncertainty measures*** designed specifically for AL document annotation selection and test them systematically, which is our ***primary contribution***. Traditional token-level uncertainty scores are inadequate for our task, where the sampling unit is an entire document. A document provides more focused context than an entire corpus with many documents, and broader context than a single sentence.

## Methods

### Active learning overview

A typical supervised learning setting requires a large amount of labeled data, which can be expensive and time-consuming to obtain. AL addresses this challenge by iteratively and strategically selecting the data points the model finds most uncertain and presenting them for HDE annotation, reducing annotation effort while maximizing the model’s learning efficiency^18^. This study aims to use an AL pipeline to automatically identify key phrases to facilitate ontology curation with minimal HDE annotation efforts.

In this paper, an AL cycle refers to the learning process between two stages, e.g., θ_x_ to θ_x+1_, where θ_x_ represents the model state after x AL cycles. Each cycle includes selecting the most informative (i.e., the most uncertain) documents, obtaining HDE annotations, and retraining the model with the newly human-labeled data (Figure 1). Our AL cycle operates as follows (Figure 1):

**Figure 1.**
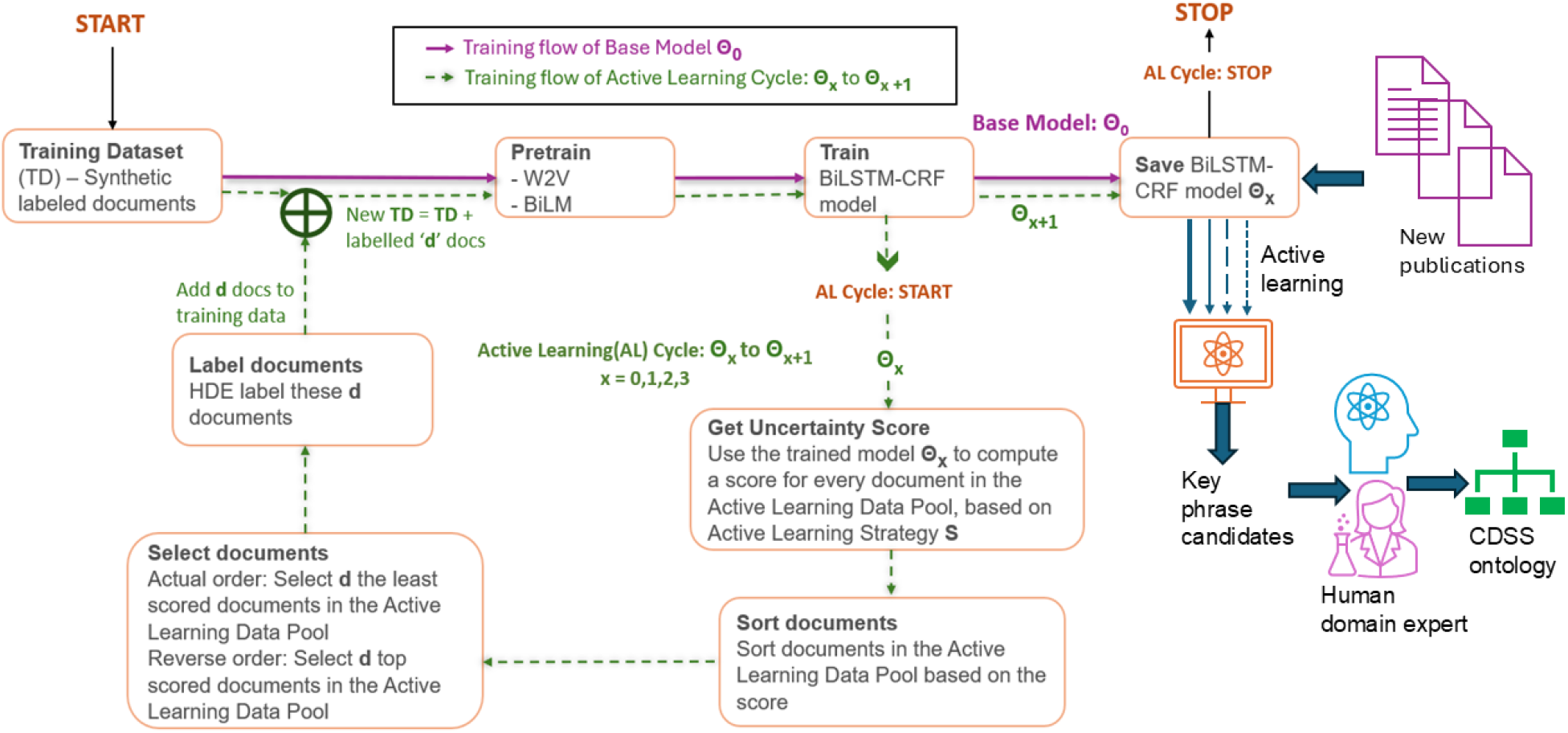
Active learning cycle for automatic key phrase identification and potential downstream applications

- At stage θ_x_, a BiLSTM-CRF model classifies tokens into five possible tags: B-KP (beginning of a key phrase), I-KP (inside a key phrase), O-KP (outside a key phrase), Start (start of the sequence), and Stop (end of the sequence). The model assigns each token a probability of belonging to one of five categories. The probabilities form the basis for later uncertainty measures and confidence score computation.
- After the confidence scores calculation, the documents are sorted based on these scores, which prioritizes the most informative (i.e., uncertain) documents for HDE annotation.
- After HDE annotation, the newly labeled documents are added to the updated training dataset and removed from the AL data pool, a critical step to avoid overfitting.
- Once the newly labeled documents are incorporated into the updated training dataset, the BiLSTM-CRF model is retrained using the updated training dataset, which is denoted as θ_x+1_. Then the next AL cycle starts. This iterative process continues until a stopping criterion (such as performance saturation) is met.

### Active learning framework

The AL framework begins with a BiLSTM-CRF model because it is lightweight, computationally efficient, well-established, interpretable, and stable for sequence labeling tasks. Our prior publication detailed the model development and comparisons^10^. Then, a pool-based AL strategy, as outlined by Settles et al.^12^, was used to iteratively enhance the model by selecting the most informative documents, allowing the model to focus on the most uncertain data points to maximize learning efficiency. Figure 2 illustrates an AL lifecycle, from θ_0_ to θ_1_.

**Figure 2.**
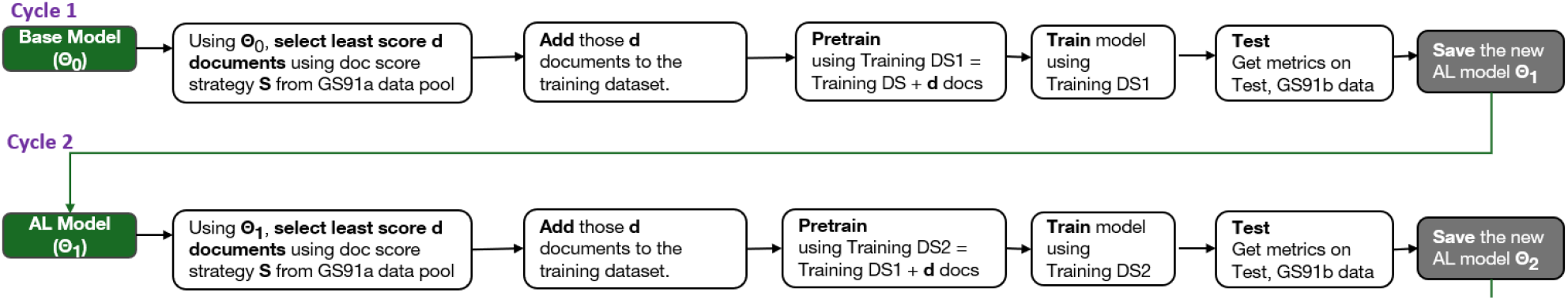
Active Learning life cycle examples from θ_0_ to θ_1_ with elaborated steps

- θ_0_: The initial model (θ_0_) is trained on 500 synthetically labeled CDSS documents^10^.
- θ_1_: Then, the model is retrained on 500 synthetic documents + 5 selected HDE annotated documents.
- θ_2_ to θ_4_ will each time add 5 new documents from the last cycle into the training dataset pool; i.e., the final cycle includes 500 synthetic documents + 20 HDE-annotated documents (15 from θ_1_, θ_2_, and θ_3_ + 5 new documents).

### Uncertainty sampling strategies

To identify documents for HDE annotation, we utilize well-established uncertainty metrics such as Minimum Token Probability (MTP), Margin, and Token Entropy. These metrics help identify tokens or sequences where the model’s predictions show the lowest confidence.

***Minimum Token Probability (MTP)***^5^ assesses uncertainty by considering the highest probability assigned to a token tag. A lower highest probability indicates greater uncertainty, as the model is less confident in any single prediction.

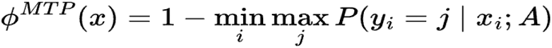

Where j ranges over all possible token tags. A is a parameter called the transition matrix. X_i_ is the i-th token in the sentence, and y_i_ is the tag that the i-th token can assume.

***Token Entropy***^19^ measures uncertainty by calculating the entropy of the probability distribution across all possible tags for each token. Greater entropy indicates less information available to the model to make a confident prediction for a given token. By selecting tokens with the greatest entropy for AL, the system focuses on uncertain areas.

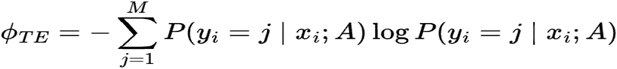

Where j ranges over all possible token tags. A is a parameter called a transfer matrix. X_i_ is the token. Y_i_ is the tag that the i-th token in the sentence can assume; M is the number of tags, in our case, it is 5.

***Margin Method***^20^ calculates uncertainty by examining the difference between the probabilities of the two most likely predicted labels for a token. A smaller margin reflects greater uncertainty, as the model finds it harder to differentiate between the top predictions. While the method still computes the full probability distribution, it utilizes only the top two probabilities, making it more computationally efficient than Token Entropy, which requires contributions from the entire distribution. The Margin Method is effective for identifying instances near prediction boundaries, where the model’s confidence is the weakest.

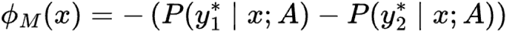

Where A is a parameter called a transfer matrix. *y*^*^ _1_ and *y*^*^ _2_ are the most and the second most likely tags for the token x, respectively.

### Document-level aggregation

We introduce novel methods—***KPSum, KPAvg, DocSum***, and ***DocAvg***—to aggregate token-level confidence scores into document-level confidence scores. Gupta used similar measures ChowSum and ChowAverage^21^, which were aggregated uncertainty at the sentence level. Our approach differs in two ways. First, document-level aggregation better aligns with our AL task, i.e., a document is the annotation unit. Second, we propose ***KPSum*** and ***KPAvg***, both aggregate uncertainties only over tokens likely to belong to key phrases, enabling more targeted and efficient sampling. This key phrase-centric perspective is absent from prior work, distinguishing our method from others.

***DocSum*** sums the model uncertainty (measured by Token Entropy, Margin, or MTP) across all tokens in a document. ***DocSum*** identifies documents with the highest overall uncertainty for HDE review, which tends to favor longer documents, as they naturally contain more tokens.

***DocAvg*** calculates the arithmetic mean uncertainty per token in the document. This method selects documents with high mean uncertainty, balancing document length and uncertainty.

***KPSum (Key Phrase Sum)*** sums the uncertainty scores of all identified key phrases (instead of all tokens) within a document. It prioritizes documents based on the collective uncertainty of the predicted key phrases. As a result, the approach can be biased toward documents with more key phrases.

***KPAvg (Key Phrase Avg)*** takes the arithmetic average uncertainty of the predicted key phrases per document, regardless of how many key phrases are predicted.

Both ***KPSum*** and ***KPAvg*** focus on the most ambiguous key phrases predicted by the BiLSTM-CRF model. These methods prioritize key phrase tokens over all tokens. Therefore, ***KPSum*** and ***KPAvg*** ensure that both the model and HDE attention focus on documents containing the most uncertain key phrases. A table summarizes the strategies presented in the full version of this paper, available on the GitHub repository (https://github.com/CDSS4PCP).

#### Document Sorting and Selection

After computing the document-level confidence scores, all documents in the AL data pool are sorted in descending order of their uncertainty.

#### Human Domain Expert Review

The most uncertain documents, as identified through the confidence scores, are reviewed and annotated by the HDE.

#### Retraining the Model

Once the documents are annotated by HDE, they will be added to the training dataset and removed from the AL data pool. The BiLSTM-CRF model is then retrained using the updated training dataset. This process enables the model to learn from continuously updated ground-truth labels. Table 1 summarizes the systematic evaluation of different measures, strategies, and their combinations. Random sampling serves as a control to provide a baseline performance.

**Table 1.**
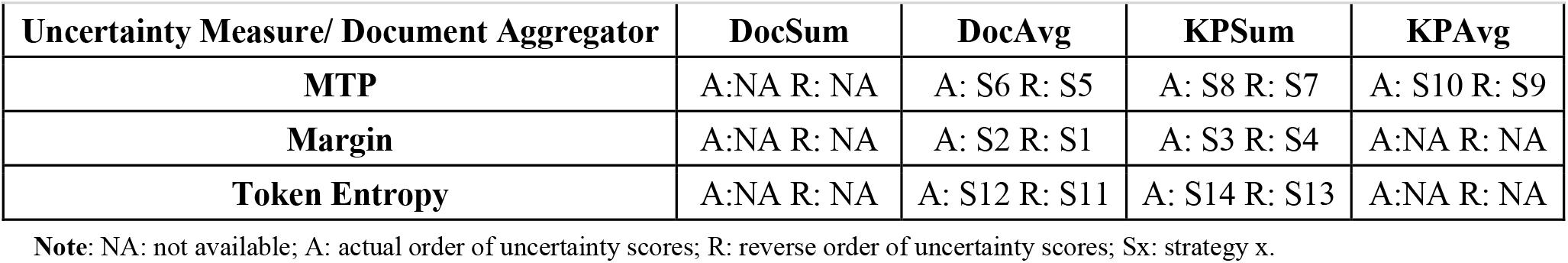
All the active learning strategies experimented within this study.

### Document order in active learning cycles

The order in which documents are selected during AL cycles may significantly shape the model’s learning trajectory. Therefore, we evaluated two strategies:

- **Actual Order**: Documents are prioritized based on their uncertainty (e.g., calculated by Token Entropy and ***KPSum***), i.e., the most uncertain documents are presented first to HDE.
- **Reverse Order**: Documents are presented in reverse order of their uncertainty.

### Experiment strategies

In each experiment, we test a specific strategy as shown in Table 1, i.e., a combination of the uncertainty measure (e.g., Margin, MTP, Token Entropy), document aggregator (e.g., ***DocSum, DocAvg, KPSum, KPAvg***), and document order (e.g., Actual, Reverse). Recall and F1 improvement were used to compare the model’s performance. The ***DocSum***-based strategies were not included in the final performance comparison, as they consistently favored longer documents. However, ***DocSum*** was used to develop the model and optimize hyperparameters.

### Dataset overview

**The Initial Training Dataset** consists of 500 documents from PubMed and annotated with synthetic labels^10^. This dataset is used to train the base model (θ_0_) before AL begins. **Active Learning Data Pool** comprises 42 documents. Five documents were selected based on model-predicted uncertainty measures in each AL cycle, and then the selected documents with the HDEs annotation (gold standard data) were added to the updated training dataset.

**The Test Set** consists of 49 HDE-labeled documents, which are untouched throughout the AL cycles. This dataset was used to evaluate the model’s performance, ensuring that improvements observed during AL are not due to data leakage but reflect the model’s genuine generalizability. The HDE-labeled documents were annotated by three medical domain experts, and the Kappa rates were between 0.73 and 0.97^10, 22, 23^. Prior publications elaborate on the annotation process^10, 22, 23^.

### Metrics used in the experiments

We used Recall, F1-score, and weighted F-measure: fbeta_score^30^ to assess model performance. These metrics are compared across AL cycles to measure the model improvement as it incorporates HDE feedback. Recall is selected as the primary metric of interest, which is motivated by the goal of minimizing the risk of missing any key phrases. F1-score provides a more holistic view of model performance by balancing precision and recall. Therefore, we prioritize recall but also consider F1-score. Weighted F-measure is used to provide additional insights. We used beta values of 5 and 10, i.e., recall is 5 times or 10 times more important than precision^30^.

### Hyperparameters and experiment settings

To ensure fair, robust comparisons across all AL strategies, we systematically tuned the following hyperparameters and compared the results. We varied one hyperparameter at a time, while holding all others constant, to assess its impact on model performance. We then combined the best-performing values.

- Number of AL cycles: 4
- Number of documents added per cycle: 5
- Number of copies per document per cycle: 1
- BiLM retrained: Yes (the Language Model, BiLM, is retrained after each AL cycle)
- Sequence length (BiLM): 10
- Learning rate: 0.001
- Dropout rate: 0.3
- Repetitions: we repeated each experiment 25 times and reported the average recall and F1 score along with their 95% confidence intervals (mean ± 1.96 × standard error).

The number of AL cycles is a key hyperparameter that influences model improvement over time. Typically, AL continues until a predefined stopping criterion is satisfied^24^. In our study, we initially experimented with eight cycles but found that most performance gains occurred early, with subsequent cycles yielding diminishing returns.

Therefore, we included four AL cycles in all experiments. Additionally, informed by our previous experiments^10^, we found that incorporating 1–2 HDE-annotated documents for every 100 synthetically labeled documents yields optimal results when HDE-labeled documents are limited. Given these considerations and 20 usable HDE-annotated documents for AL, we settled on four cycles and added five documents per cycle. Of the 42 HDE-labeled documents in our AL data pool, we allocated 20 for AL and reserved the remaining 22 to maintain variability in document selection across strategies. This was a critical design choice—if we had used all 42 documents for each strategy, the experiments would have become deterministic, limiting our ability to compare document selection and its effect on model performance. By limiting each strategy to 20 documents, we ensured diversity in selections, enabling fairer and more informative comparisons across AL strategies.

## Results

### Evaluation results by different active learning strategies

The random sampling results serve as a baseline for evaluating the effectiveness of the AL strategies. Random sampling results show an initial improvement in both recall and F1 (β = 1) from θ_0_ to θ_2_, with recall peaking at θ_2_ (0.587) before declining (Table 2). This fluctuation suggests inconsistent sampling quality, and the lack of targeted document selection likely limits model improvement beyond the early rounds. F1 (β = 1) remains relatively stable after θ_1_, indicating general knowledge gain but lower efficiency compared to uncertainty-based strategies. Figure 3 (Token Entropy) presents the average Recall and F1-score metrics for each AL cycle and strategy (detailed in Table 1), as well as comparisons between random sampling and ***KPSum*** actual order. The complete set of MTP and Margin results is available in the GitHub repository (https://github.com/CDSS4PCP).

**Table 2.** Performance of recall and F-1 for the random sampling across four active learning cycles (θ_0_ – θ_4_)

| Random (arithmetic average ± 1.96 standard error; $n = 25$) | | | | |
| --- | --- | --- | --- | --- |
| | Recall AVG | F1 AVG ($\beta = 1$) | Weighted F1 AVG ($\beta = 5$) | Weighted F1 AVG ($\beta = 10$) |
| $\theta_0$ | $0.440 \pm 0.0135$ | $0.543 \pm 0.0098$ | $0.446 \pm 0.0133$ | $0.441 \pm 0.0134$ |
| $\theta_1$ | $0.523 \pm 0.0135$ | **$0.579 \pm 0.0057$** | $0.527 \pm 0.0129$ | $0.524 \pm 0.0134$ |
| $\theta_2$ | **$0.587 \pm 0.0108$** | $0.577 \pm 0.0047$ | **$0.586 \pm 0.0099$** | **$0.587 \pm 0.0106$** |
| $\theta_3$ | $0.519 \pm 0.0135$ | $0.571 \pm 0.0088$ | $0.523 \pm 0.0127$ | $0.520 \pm 0.0130$ |
| $\theta_4$ | $0.509 \pm 0.0172$ | $0.574 \pm 0.0112$ | $0.514 \pm 0.0168$ | $0.510 \pm 0.0171$ |

**Figure 3.**
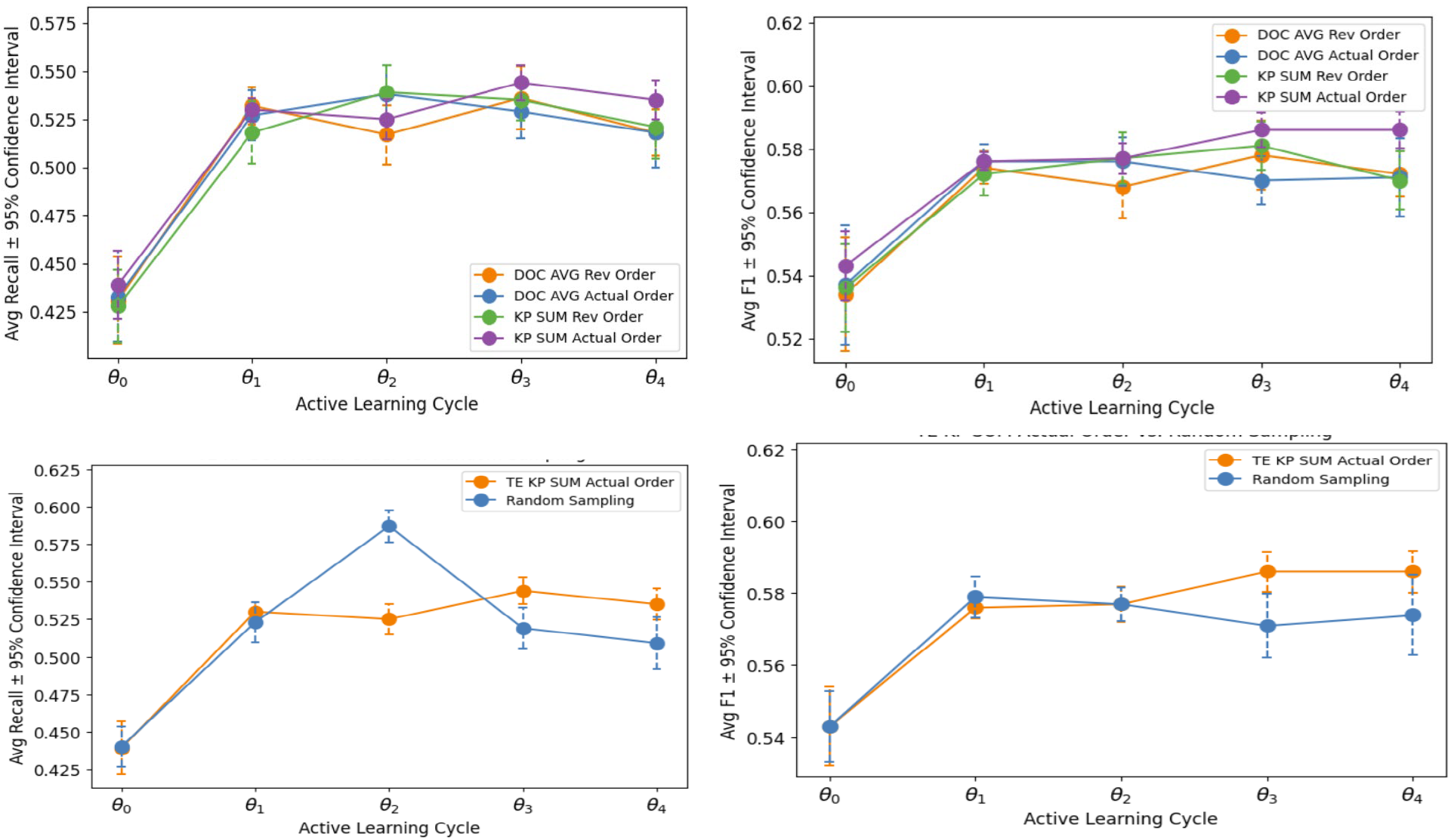
Recall (upper left) and F1 score (upper right) progress across AL cycles for Token Entropy (TE); average recall (lower left) and F1 (lower right) comparison of TE ***KPSum*** actual and random sampling; the results are based on 25 times of experiments

Across all four Token Entropy strategies (Figure 3), ***DocAvg*** and ***KPSum*** present consistent improvements in both recall and F1 over the AL cycles. The ***DocAvg*** actual order and ***KPSum*** actual order strategies show slightly higher final F1 scores (0.586) by θ_3_, suggesting better overall generalization. ***KPSum*** reverse order shows smaller F1 gains than the actual-order strategies. Overall, the trends indicate that actual order sampling tends to be slightly more effective than reverse order. ***DocAvg*** and ***KPSum*** yield comparable performance improvements over time.

### Evaluation results comparison across strategies

We noticed that most performance improvement occurs between the initial model (θ_0_) and the first cycle (θ_1_) when comparing Token Entropy evaluation results across strategies. In later cycles, recall fluctuates, suggesting performance gains begin to plateau. F1 scores show similar trends to the recall. Our results show a slightly larger improvement in recall than in F1 scores between θ_1_ and θ_0_. When we compared average recall and F1 Scores across AL cycles for the Token Entropy KPSum Actual strategy versus random sampling, we noticed that random sampling tends to produce early but less stable gains, while Active Learning via ***KPSum*** Actual leads to more gradual but sustained improvements in both recall and F1 scores. The results for Margin and MTP strategies show similar results to Token Entropy.

### Original and weighted F1 values

Table 3 reports the weighted F1 scores (± 1.96 standard error) for Token Entropy–based AL strategies evaluated at β = 1, β = 5, and β = 10 across five AL cycles (θ_0_ to θ_4_). At θ_0_, all strategies start with comparable scores, ranging from 0.430 to 0.445. ***DocAvg*** Actual Order shows consistent performance, reaching 0.542 (β = 5) at θ_2_. ***KPSum*** Actual Order attains the highest score of 0.546 (β = 5) at θ_2_, while also maintaining stable results across other cycles. ***KPSum*** Reverse Order peaks at θ_2_ with 0.542 (β = 5) and slightly declines in later cycles. ***DocAvg*** Reverse Order sees a rise from 0.439 (θ_0_, β = 5) to 0.541 (θ_2_, β = 5), then slightly fluctuates. Across all strategies and cycles, the β = 5 and β = 10 values remain closely aligned, differing only marginally within each configuration. The weighted F1 scores for Random sampling are included in Table 2. The results suggest that while random sampling provides early performance gains, it lacks consistency in maintaining or improving performance in later stages of the active learning process. The weighted F1 scores for Margin and MTP are available via a GitHub repository in the full version of this paper (https://github.com/CDSS4PCP).

**Table 3.**
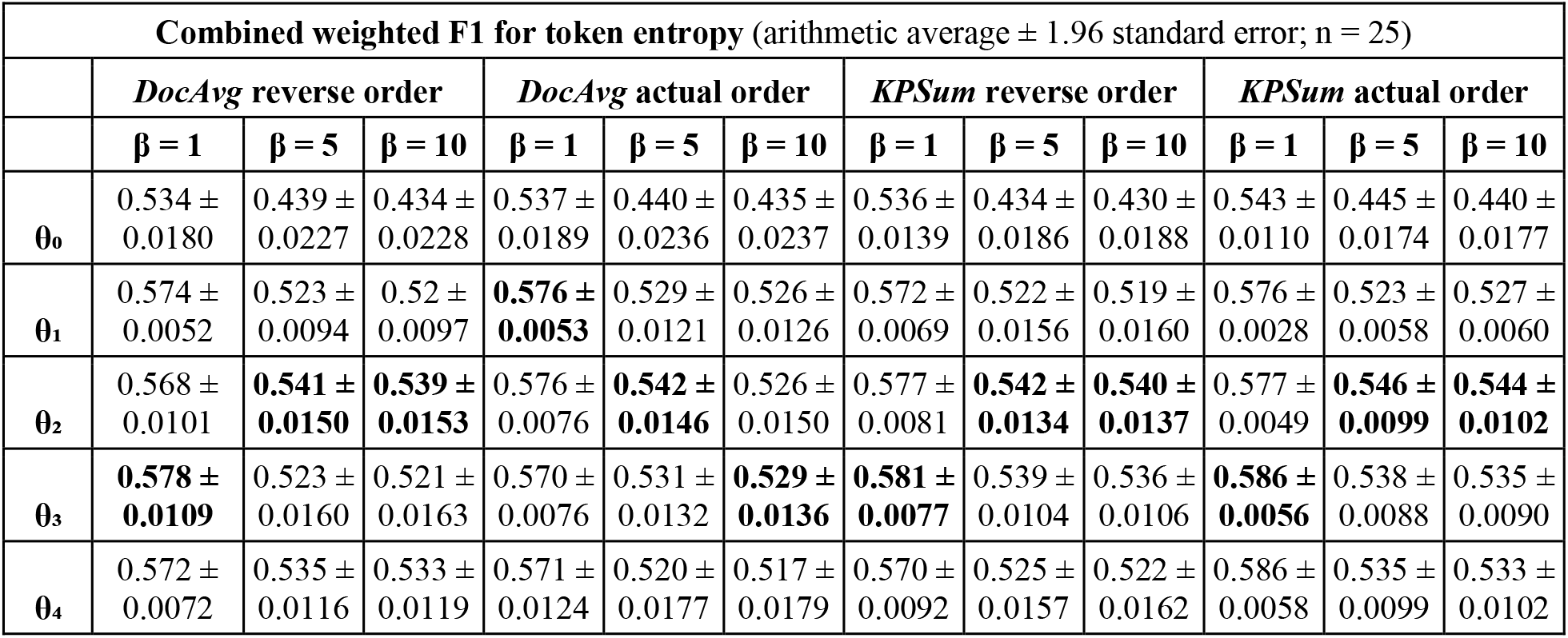
Weighted F1 scores for token entropy-based active learning strategies, *β* = 1, *β* = 5, AND *β* = 10.

| Combined weighted F1 for token entropy (arithmetic average $\pm 1.96$ standard error; $n = 25$) | | | | | | | | | | | | |
| --- | --- | --- | --- | --- | --- | --- | --- | --- | --- | --- | --- | --- |
|  | **DocAvg reverse order** |  |  | **DocAvg actual order** |  |  | **KPSum reverse order** |  |  | **KPSum actual order** |  |  |
| | $\beta = 1$ | $\beta = 5$ | $\beta = 10$ | $\beta = 1$ | $\beta = 5$ | $\beta = 10$ | $\beta = 1$ | $\beta = 5$ | $\beta = 10$ | $\beta = 1$ | $\beta = 5$ | $\beta = 10$ |
| $\theta_0$ | 0.534 ± 0.0180 | 0.439 ± 0.0227 | 0.434 ± 0.0228 | 0.537 ± 0.0189 | 0.440 ± 0.0236 | 0.435 ± 0.0237 | 0.536 ± 0.0139 | 0.434 ± 0.0186 | 0.430 ± 0.0188 | 0.543 ± 0.0110 | 0.445 ± 0.0174 | 0.440 ± 0.0177 |
| $\theta_1$ | 0.574 ± 0.0052 | 0.523 ± 0.0094 | 0.52 ± 0.0097 | **0.576 ± 0.0053** | 0.529 ± 0.0121 | 0.526 ± 0.0126 | 0.572 ± 0.0069 | 0.522 ± 0.0156 | 0.519 ± 0.0160 | 0.576 ± 0.0028 | 0.523 ± 0.0058 | 0.527 ± 0.0060 |
| $\theta_2$ | 0.568 ± 0.0101 | **0.541 ± 0.0150** | **0.539 ± 0.0153** | 0.576 ± 0.0076 | **0.542 ± 0.0146** | 0.526 ± 0.0150 | 0.577 ± 0.0081 | **0.542 ± 0.0134** | **0.540 ± 0.0137** | 0.577 ± 0.0049 | **0.546 ± 0.0099** | **0.544 ± 0.0102** |
| $\theta_3$ | **0.578 ± 0.0109** | 0.523 ± 0.0160 | 0.521 ± 0.0163 | 0.570 ± 0.0076 | 0.531 ± 0.0132 | **0.529 ± 0.0136** | **0.581 ± 0.0077** | 0.539 ± 0.0104 | 0.536 ± 0.0106 | **0.586 ± 0.0056** | 0.538 ± 0.0088 | 0.535 ± 0.0090 |
| $\theta_4$ | 0.572 ± 0.0072 | 0.535 ± 0.0116 | 0.533 ± 0.0119 | 0.571 ± 0.0124 | 0.520 ± 0.0177 | 0.517 ± 0.0179 | 0.570 ± 0.0092 | 0.525 ± 0.0157 | 0.522 ± 0.0162 | 0.586 ± 0.0058 | 0.535 ± 0.0099 | 0.533 ± 0.0102 |

## Discussion

### Significance and practical implications of the work

The study’s primary contribution is proposing new ***document-level uncertainty sampling measures***, distinct from existing token-level and sentence-level measures. We prioritize key phrases rather than all tokens in ***KPSum*** and ***KPAvg***, which distinguishes our work from others. We propose these new measures to leverage document-level context during AL model training and learning, which is richer than token-level context but still more focused than sentence-level context from a larger corpus. The systematic comparison results demonstrate the usage of document-level measures in AL model performance. However, the differences among token-level, sentence-level, and document-level measures still require a carefully designed study to demonstrate, which can be a future direction. The newly proposed document-level measures are excellent complements for selecting documents for annotation during AL. Our results indicate that ***KPSum*** and ***DocAvg*** are promising for informing document selection decisions, making model learning more informative and efficient. Given our use case, we anticipate this study will open a new venue for research on automating ontology curation in the real world. The actual annotation efficiency and corresponding improvements from these new measures require another carefully designed future study to demonstrate. However, we believe the new document-level measures are important additions to make the annotation process more targeted, streamlined, and efficient. Realistically, ontology curation needs automation to make the process streamlined and sustainable. The AL pipeline and the document-level measures are promising attempts to assist the process, although performance has room to improve before the model can be adopted in real-world use.

### The critical role of HDE annotated documents in model learning

Most performance improvement occurs during the transition from θ_0_ to θ_1_, regardless of the strategy, which aligns with others’ findings^25^. This highlights the importance of the initial rounds of HDE feedback, as they address the most critical gaps in the model’s learning and drive rapid improvements. It emphasizes the need to prioritize high-quality HDE labels during the initial cycles to maximize their impact on the AL process. Even when documents are added randomly, there is still a significant improvement from θ_0_ to θ_1_ in both F1 and recall, indicating that HDE-labeled data are a major factor in the performance boost. Another unique characteristic of our use case is the task’s complexity, which differs significantly from a typical binary predictive task. The three HDEs could not reach 100% consensus even after discussions during HDE annotation, showing the task is challenging for the model.

### Identifying saturation in active learning cycles

Determining an effective stopping criterion is crucial for AL with iterative cycles and ensures effective use of computing resources. A well-defined stopping condition prevents excessive labeling costs and mitigates the risk of overfitting. One approach is to monitor the convergence of learning by evaluating the generalization error on a test dataset, as discussed by H Ishibashi, et al^24^. However, obtaining a sufficiently large and representative test set may not always be feasible. A practical alternative is to define saturation-based stopping criteria, where the performance improvements fall below a predefined threshold or when key evaluation metrics begin to fluctuate around a stable value. In Figure 3, recall generally increases from θ_0_ to θ_2_, after which it either plateaus or fluctuates slightly without consistent gains. F1 scores show a similar trend. However, we acknowledge that we may observe earlier saturation in our experiments due to the limited size of our AL data pool.

### Impact of KPSum strategies on recall and F1 improvement in active learning

We note that ***KPSum*** Actual presents the most consistent and stable improvements in recall, with the highest recall gains and lower variation in the confidence intervals, indicating more stable learning. ***KPSum*** calculates document uncertainty solely based on key phrases, which favors documents with more key phrases. Although the per-cycle recall gain of ***KPSum*** Actual may not always be dramatically higher than other strategies, its sustained improvement and model stability suggest a long-term advantage.

### F1 and recall dynamics in KPAvg strategies: Actual vs. Reverse

***KPAvg*** Reverse consistently achieves higher F1 scores than ***KPAvg*** Actual across cycles, indicating that selecting documents with lower key phrase uncertainty (as in Reverse) helps the model reduce false positives, improving precision and overall F1. Although ***KPAvg*** Actual was expected to improve recall by selecting documents with higher key phrase uncertainty, the recall improvements are marginal and inconsistent compared to the Reverse strategy.

### Active learning vs. random selection: impact on recall and model stability

We noted that both the AL (***KPSum*** Actual) and Random selection strategies improve model performance; the difference lies in the stability of these improvements. ***KPSum*** Actual shows more reliable gains, especially after the initial learning cycles; i.e., the F1 and Recall scores show a more consistent upward trend than Random sampling. In contrast, Random selection improves initially but remains volatile. After a certain point, recall and F1 scores for Random selection can decline, highlighting its inefficiency in refining model performance. These results align with findings by Ray, et al^25^. Our results further underscore the value of a structured, uncertainty-based approach for document selection, which offers more reliable and stable performance gains than Random selection.

### F1 scores and weighted F1 scores

F-measures are typically used in machine learning, information retrieval, and NLP tasks. Although recall is more important in our use case, we still report both original and weighted F-measures to provide a more balanced view of model performance. From Table 3, we noticed weighted F1 scores (β = 5, 10) show a similar trend. Weighted F1 values were approximately 0.03 to 0.05 lower than the original F1 values across strategies. Moreover, weighted F1 values peaked at θ_2_ consistently across most strategies, including random sampling. The original F1 value peaked inconsistently, including θ_1_, θ_2_, and θ_3_. Overall, weighted F1 values provide important complementary measures to original F1 values and recalls.

### How could the results be used?

We initiated the investigation to leverage automatic key phrase identification to support long-term maintenance of the CDSS ontology. The results presented in this paper show promise but modest gains in model performance; therefore, the model is not yet ready for practical use. Given the complexity of our task and the long-term maintenance challenges, the initial promising results are encouraging, even though the model is not yet ready for practical use. This is also reflected in work using LLMs to extract semantics^15^, which used the most advanced technology to achieve similar goals; however, it achieved results comparable to ours. This indicates that the task is indeed challenging and there is room for improvement. Including HDE in the loop is necessary in the foreseeable future. Meanwhile, exploring ways to improve model performance remains a priority.

### Other considerations

Due to space limits, we can only include Token Entropy results in this paper. However, Margin and Minimum Token Probability show similar results and trends as Token Entropy. This study builds on the prior work^10^. We reported the performance of our AL pipeline versus the transformer-based model in a prior paper^10^, along with an ablation study of the BiLSTM model^10^. We hypothesized that Reverse order would yield quicker improvements in the early stages (θ_0_ to θ_1_) and that Actual order would show more sustained improvements later (θ_0_ to θ_2_ or θ_3_). This is why we experimented with both methods. However, the observed pattern did not hold across all strategies and showed advantages in some, possibly due to the limited dataset. While the Reverse order showed quicker initial gains, the Actual order did not consistently lead to greater long-term improvements as expected.

### Limitations and future work

Our study has several limitations: (1) the performance of the current AL pipeline can be improved further to be applied in real-world operations. (2) In this study, we have a limited number of ground truth documents. With more ground truth documents, researchers may obtain different results. However, we believe the approach described in this paper is still valid and can be reused. (3) In this study, we explored basic aggregation strategies for estimating document-level uncertainty, primarily focusing on sum-based (e.g., ***KPSum, DocSum***) and average-based (e.g., ***KPAvg, DocAvg***) methods. Future work may include advanced mathematical computations, such as exponential or logarithmic calculations. Future work could also explore alternative aggregation techniques, such as trimmed mean or weighted mean. Techniques such as median, mode, or box-plot-based selection (e.g., interquartile range filtering) may also yield more robust document representations, especially in skewed distributions.

In addition, the real impact of document-level uncertainty sampling in HDE annotation tasks can be measured based on the effort saved over sampling algorithms^26-29^. Although prior studies^31,32^ have examined token-level uncertainty in NER and uncertainty-based instance selection more broadly, our work extends these efforts by proposing task-specific document-level aggregation techniques for AL in a key phrase identification task. This represents a novel abstraction layer between token uncertainty and document selection, which can be especially important in ontology construction and maintenance, where annotation decisions occur at the document level. In addition, the methodology described in this paper is not specific to BiLSTM-CRF model. Future work could apply the same approach to other types of language models.

## Conclusion

We proposed new document-level measures for uncertainty sampling in an AL framework (based on a BiLSTM-CRF model) to identify candidate key phrases for a CDSS ontology and evaluated them systematically. A key contribution is the novel document-level uncertainty aggregation strategies (***KPSum, KPAvg, DocSum***, and ***DocAvg***), which enable more effective, transparent, and reproducible document selection for HDE annotation. The experimental results show ***KPSum*** (Actual) is the most effective strategy for key phrase identification in our use case. This study offers practical benchmarks for future research through systematic comparison of various uncertainty measures. These methods can be adapted to select optimal approaches and measures for other sequence labeling tasks and datasets. Overall, this work contributes to the underexplored application of AL in deep learning-based key phrase identification, providing a scalable and adaptive foundation for efficient HDE annotation in NLP.

## Data Availability

The full set of evaluation results and a full version of the manuscript can be found in the link: https://github.com/CDSS4PCP/NLP_KPIdentify/tree/active_learning_files/ActiveLearningStrategiesResults

## Acknowledgment

This study is funded by the National Institute of General Medical Sciences (R01GM138589) at the National Institutes of Health and partially through P20GM121342 of NIGMS and partially through # OIA-2242812 of the NSF.

